# Genetic similarity among 178 disease phenotypes predicts therapeutic and side effects for 1,711 drugs

**DOI:** 10.1101/2025.05.13.25327511

**Authors:** Panagiotis N. Lalagkas, Rachel D. Melamed

**Affiliations:** Department of Biological Sciences, University of Massachusetts Lowell, Lowell MA, USA

## Abstract

Human genetics demonstrates great potential for drug discovery, but challenges in identifying causal genes limit its clinical translation. Pleiotropy, the phenomenon where genetic variants or genes influence multiple traits, has been previously used to identify drug targets shared between phenotypically similar diseases. Here, we expand the use of pleiotropy to develop and evaluate a gene-agnostic method that predicts novel drug therapeutic and side effects across the phenome. We hypothesize that diseases with high genetic similarity to a drug’s known indications can point to new drug uses. To test this, we develop five metrics to quantify the genetic similarity between pairs of 178 diseases integrating genome-wide genetic correlation, gene-level associations and tissue-specific gene regulation. Comparing these metrics with data on indications and side effects of 1,711 common drugs, we find that more genetically similar diseases tend to share more drugs. We then use genetic similarity to predict drug therapeutic effects: our predictions with probability >0.1 show a 2.03-fold increased likelihood of progressing from Phase I clinical trials to regulatory approval. As well, predictions with probability >0.2 are 1.42 times more likely to correspond to true adverse effects. Notably, the indications model predicts side effects better than expected by chance, and vice-versa, implying a shared genetic basis for therapeutic and adverse drug effects. Together, our results suggest that genetic similarity can reveal new drug-disease links, putting forward a new use of genetics that bypasses the need for disease and drug target identification.

## Introduction

Human genetics is increasingly used to identify effective drug gene targets for diseases through genome-wide association studies (GWAS) (1,2). Notably, drugs targeting genes pointed by GWAS disease-associated variants show higher success rates in clinical trials than those lacking such genetic support (3–5). Recently, this signal has been shown to predict drug side effects as well (6). However, it is challenging to link variants to causal genes that could be potential drug targets, and this approach must conservatively discard a substantial fraction of the genetic results (7,8). Therefore, there is a need for methods that can leverage the wealth of existing genetic data to accelerate drug discovery.

Pleiotropy, the phenomenon where genetic variants or genes influence multiple traits through same or distinct biological processes, is widespread in the human genome, encompassing 90% of all GWAS trait-associated loci (9–11). This shared genetic risk has been shown to explain clinical co-occurrence of diseases (12,13). Additionally, pleiotropic effects of drug gene-targets have been previously used to guide drug repurposing. Woodward et al. review methods that identify gene-targets of antipsychotic drugs to be shared between pairs of psychiatric disorders, uncovering potential drug repurposing opportunities (14). A recent study used evidence of genetic similarity between Mendelian and complex diseases to show that Mendelian disease causal genes are potential drug targets for genetically similar complex diseases (15), even if those gene-targets have not been linked to the complex disease, and even if the diseases appear unrelated phenotypically (16,17). Other work showed that shared genetics underlying clinical co-occurrence of breast cancer and complex diseases can inform drug repurposing (18). Building on these findings, here we propose a broader hypothesis: that genetic similarity between a pair of diseases can point to novel effects of drugs that are known to act on only one of the two diseases. We test this hypothesis both in terms of therapeutic and side drug effects.

## Results

### Genetically similar diseases share more drugs

We use publicly available data and develop five metrics to quantify the genetic similarity between 178 diseases (**Fig 1** shows an overview of the study framework). Each metric captures different aspects of the human genetic architecture including genome-wide genetic correlation, gene-level associations, tissue-specific gene regulation, and molecular QTL colocalization (see Methods). Consistent with prior studies, we find that diseases affecting the same body system (such as myocardial infarction and hypertension; both cardiovascular) have higher genetic similarity scores than those affecting different body systems (such as Alzheimer’s disease (neurological) and ulcerative colitis (gastrointestinal)) (p<0.05, Wilcoxon rank-sum test, two-sided; **S1A-E Figs**) (19). We define two types of drug similarity: 1) the overlap of drugs indicated for both diseases and 2) the overlap of drugs causing both diseases as side effects.

**Fig 1.**
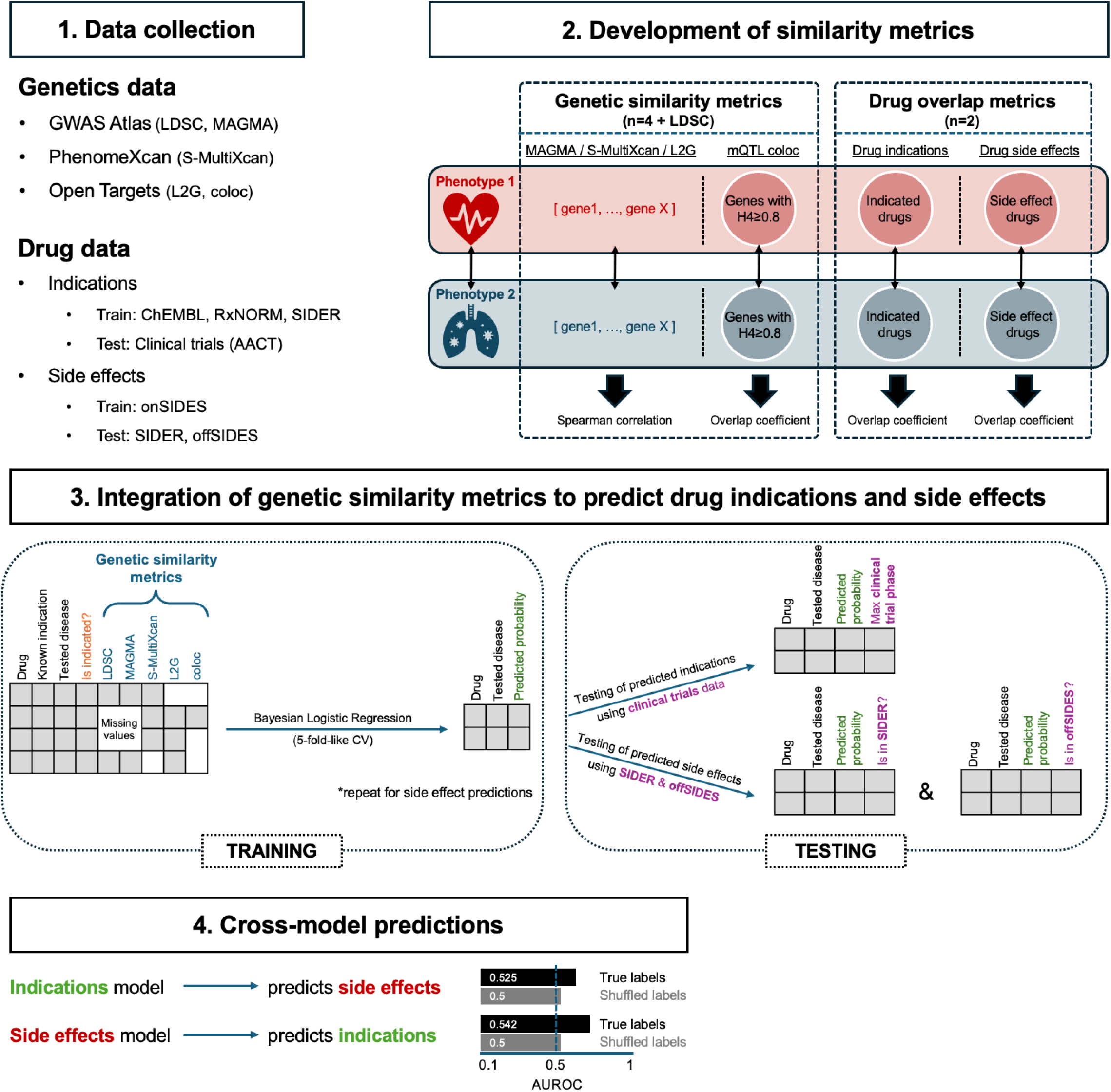
Overview of the study framework. Step 1: data collection. Step 2: construction of genetic similarity and drug-overlap metrics. Step 3: analytical workflow for integrating multiple genetic similarity metrics to predict drug-disease indications or side effects. Step 4: assessment of shared genetic architecture between indications and side effects.

After harmonizing drug and disease identifiers across multiple sources, we have 1,643 drugs indicated for 159 diseases and 1,140 drugs causing 149 diseases as side effects (**Table 1**). As expected, we find that diseases from the same body system share significantly more indicated drugs than those from different body systems, and a similar pattern is observed for side effect drugs (p<0.05, Wilcoxon rank-sum test, two-sided; **S1F-G Figs**) (20).

**Table 1.**
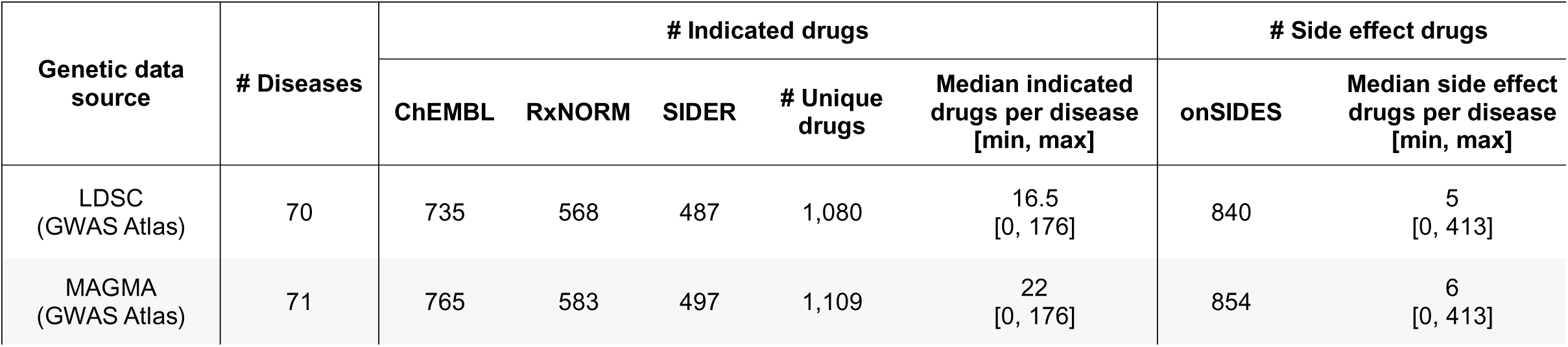
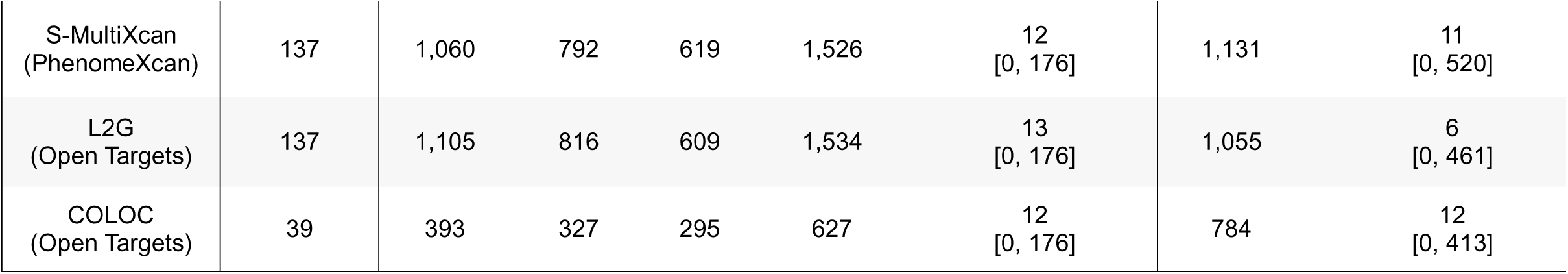
Summary of the genetic and pharmacological data used.

| Genetic data source | # Diseases | # Indicated drugs |  |  |  |  | # Side effect drugs |  |
| --- | --- | --- | --- | --- | --- | --- | --- | --- |
|  |  | ChEMBL | RxNORM | SIDER | # Unique drugs | Median indicated drugs per disease [min, max] | onSIDES | Median side effect drugs per disease [min, max] |
| LDSC (GWAS Atlas) | 70 | 735 | 568 | 487 | 1,080 | 16.5 [0, 176] | 840 | 5 [0, 413] |
| MAGMA (GWAS Atlas) | 71 | 765 | 583 | 497 | 1,109 | 22 [0, 176] | 854 | 6 [0, 413] |
| S-MultiXcan<br>(PhenomeXcan) | 137 | 1,060 | 792 | 619 | 1,526 | 12<br>[0, 176] | 1,131 | 11<br>[0, 520] |
| L2G<br>(Open Targets) | 137 | 1,105 | 816 | 609 | 1,534 | 13<br>[0, 176] | 1,055 | 6<br>[0, 461] |
| COLOC<br>(Open Targets) | 39 | 393 | 327 | 295 | 627 | 12<br>[0, 176] | 784 | 12<br>[0, 413] |

Next, we test whether pairs of diseases with higher genetic similarity have greater drug similarity. A confounding factor here is phenotypic similarity: pairs of diseases with more similar etiology, often affecting the same body system, share genetics and more drugs (21). While phenotypic similarity is well known to present opportunities for repurposing drugs for diseases with shared features, our goal is to identify the distinct potential of genetic similarity to identify drug indications that would not be obvious based only on phenotypic similarity. To isolate the signal from genetic similarity, we first stratify our analysis by annotated body system. Across nearly all genetic similarity metrics, we observe a positive correlation between genetic and drug similarity, even when analysis is restricted to diseases from different body systems (**Figure S2**). To stringently focus only on genetic similarity among phenotypically dissimilar diseases, we also use disease embeddings from the ClinGraph knowledge graph model that summarize the phenotypic similarity between pairs of diseases (**S3 Fig**) (22). We then apply a range of thresholds for identifying pairs of diseases without phenotypic similarity. Even among pairs with low phenotypic similarity, genetic similarity remains associated with drug similarity (**Fig 2**). While this pattern holds for both drug indications and side effects, the strength of association varies across genetic similarity metrics and drug similarity types. Particularly, the L2G-based metric shows weak or non-significant association with drug similarity, especially for drug side effect similarity (**Fig 2, S2 Fig**). This may be a result of the conservative design of L2G prioritizing causal genes only at genome-wide significant GWAS loci, resulting in sparser data for assessing genetic similarity (23). These results support our overall hypothesis that genetics holds useful signal for understanding drug-disease connections beyond the strongest causal genes.

**Fig 2.**
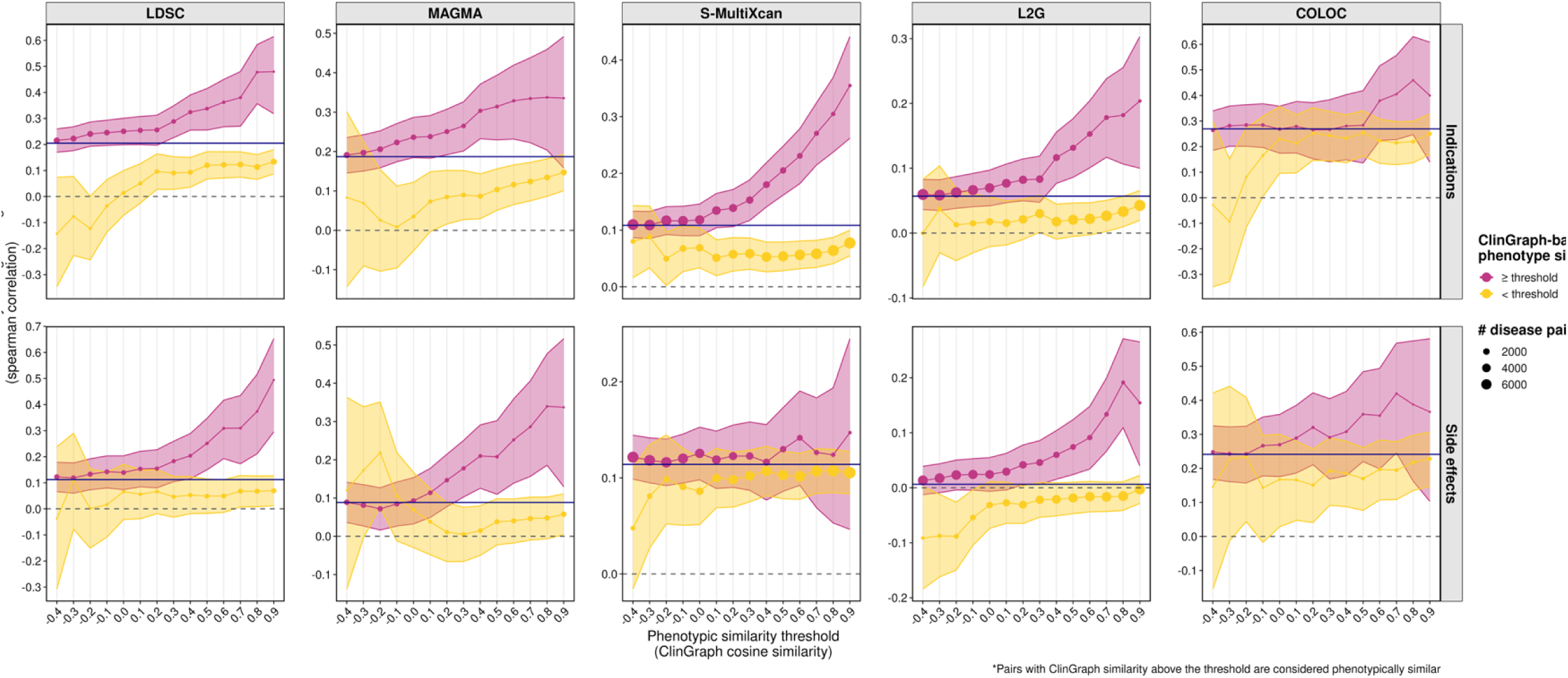
Diseases with higher genetic similarity scores share more drugs even after stratification by phenotypic similarity. Each point represents the spearman correlation between a genetic similarity metric (columns) and drug sharing across disease pairs (top row: sharing of drug indications; bottom row: sharing of drug side effects). The size of the points is proportional to the number of disease pairs contributing to each estimate. Shaded areas denote the 95% confidence interval of the observed correlation (estimated by z-transformation; R package DescTools, function SpearmanRho()). Colors denote disease pairs subsets defined by phenotypic similarity (varying x-axis threshold), defined as ClinGraph embedding cosine similarity between two diseases. The blue solid horizontal line indicates the estimated correlation using all disease pairs (no phenotypic similarity threshold applied). The red color indicates disease pairs with phenotypic similarity greater than or equal to a x-axis threshold. The yellow color indicates disease pairs with phenotypic similarity below a threshold.

### Genetic similarity distinguishes known drug indications and side effects

As we wish to build a model that can predict drug-indication pairs based on genetic similarity, we next test whether genetic similarity can distinguish a drug’s known indications from other diseases not known to be treated with that drug. For each drug-disease pair and for each genetic similarity metric, we find the greatest genetic similarity score between that disease and any of the drug’s known indications. We use the maximum similarity based on the assumption that if a disease is genetically similar to any existing drug indication, it may be a promising candidate for repurposing. Using all drug indication-disease pairs, we find that diseases known to be treated by the drug are genetically more similar to the drug’s other indications (p<0.05, Wilcoxon rank-sum test, two-sided) (**Fig 3**). This relationship holds even after analyzing disjoint sets of drug indication-disease pairs affecting the same or different body systems (**S4 Fig**) or using different thresholds of phenotypic similarity (**S5 Fig**). We observe similar results for drug side effects (**Fig 3**), which also persist after stratification by body system (**S6 Fig**) or phenotypic similarity (**S7 Fig**). Together, these findings support the utility of shared genetic architecture as a predictor for drug indication and side effects.

**Fig 3.**
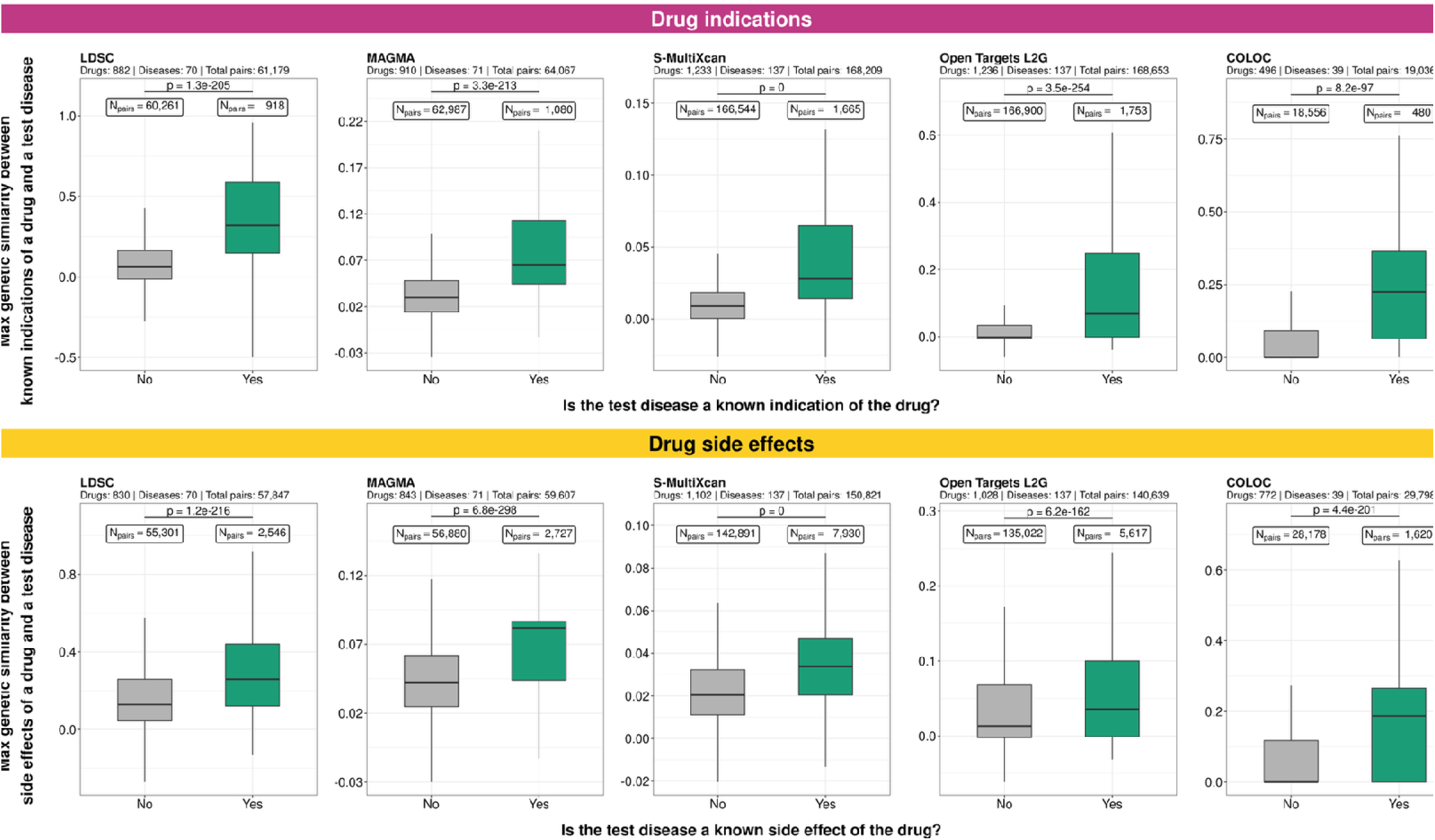
Genetic similarity distinguishes known drug indications and side effects. Top-panel: drug indications. Bottom-panel: drug side effects. Y-axis: maximum genetic similarity between a drug’s known indications (or side effects) and a tested disease. Color denotes whether the disease is a known indication (or side effect) for the drug (green) or not (gray). Outliers, estimated as values below Q1 − 1.5×IQR or above Q3 + 1.5×IQR, are not shown but included in the statistical analysis (Wilcoxon rank-sum test, two-sided). IQR: Inter-quartile range; Q1: first quartile (25th percentile); Q3: third quartile (75th percentile)

### Integrating multiple measures of genetic similarity in a predictive model for drug indications

Building on the above findings, we next integrate our measures of genetic similarity in a unified model to predict the probability that a given drug-disease pair represents a potential therapeutic indication. As above, we predict that diseases highly genetically similar to known indications of a drug are promising drug indications. But, as not all genetic similarity measures are available for all pairs of a drug indication and disease, we develop a Bayesian logistic regression model that allows us to make predictions for any drug-disease pair using all available genetic similarity data. This strategy accommodates missing information on genetic similarity of a disease to some indications of a drug, while leveraging complementary information across genetic similarity metrics.

We assess the model performance for predicting drug therapeutic effects with and without removing phenotypically similar disease pairs. Without removing phenotypically similar indications, our model is able to predict indications for held-out drug-disease pairs with an AUROC of 0.747. To disentangle genetic similarity from confounding phenotypic similarity, as above, we remove drug indication-disease pairs with phenotypic similarity (see Methods) and re-train the model. Although this approach removes genetic similarity that overlaps with phenotypic similarity, excluding informative signal, our model is still predictive of known indications (AUROC=0.559, individual disease AUROCs shown in **Fig 4A**). We note that fewer diseases are included in this analysis because the exclusion of phenotypically similar drug indication-disease pairs result in certain diseases having no indicated drugs remaining in the test set.

**Fig 4.**
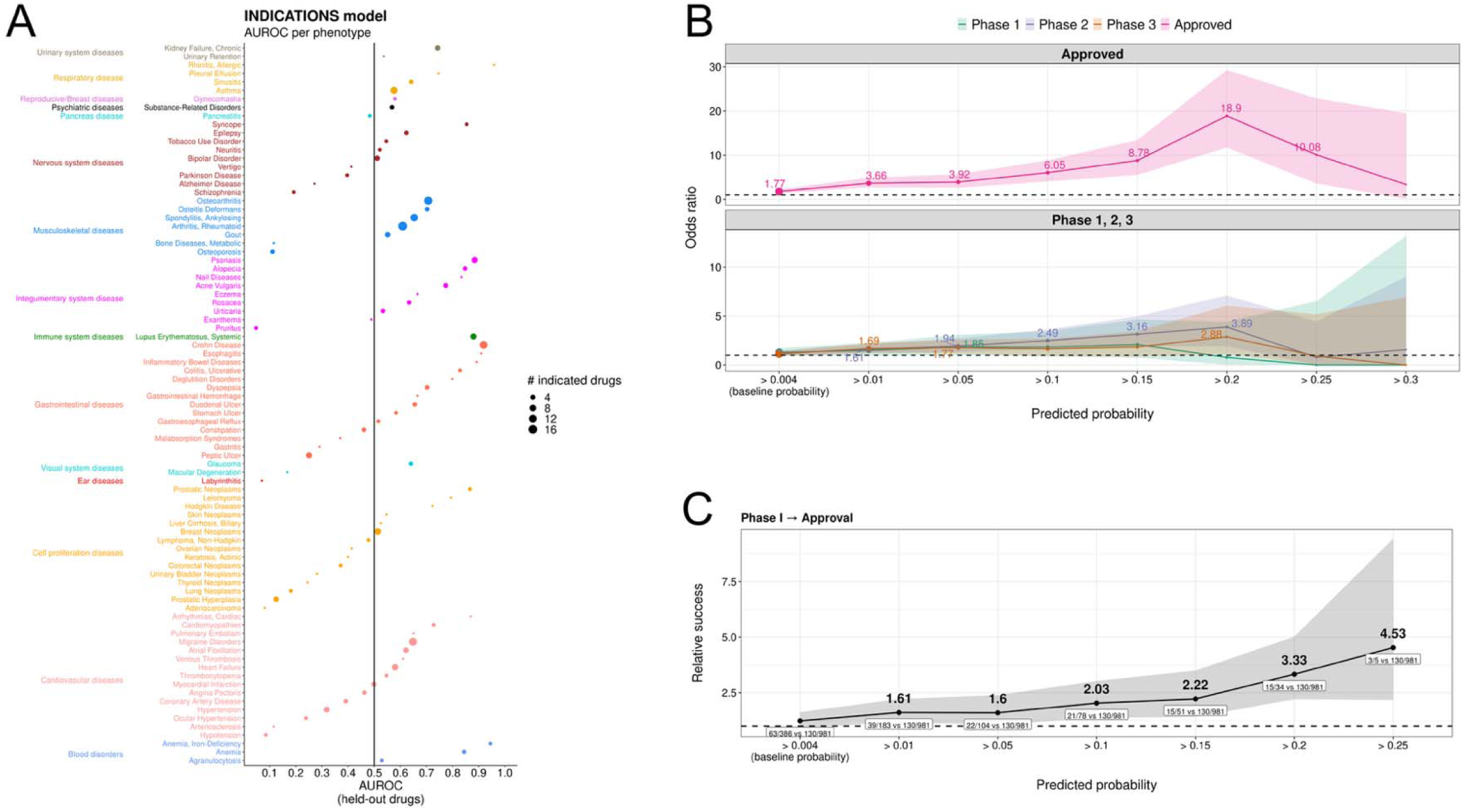
Evaluation of the indications model. **A.** AUROC (x-axis) for each tested disease (y-axis). Diseases are colored by body system (Open Targets labels). **B.** Odds ratio (y-axis) for drug-disease pairs with predicted probability above a given threshold (x-axis) compared to pairs below the baseline probability of being an indication in the test set (0.004), indicating enrichment for testing in a clinical trial phase (color) versus never tested in clinical trials. Shaded regions indicate 95% confidence intervals calculated by Fisher’s exact test (two-sided). Numbers above points show the calculated odds ratio (only when p-value<0.05). The size of the point is proportional to the number of pairs with predicted probability exceeding the threshold (x-axis). In total, 66,395 drug-disease pairs are included in this analysis (124 pairs tested in clinical trials but lacking phase information are excluded). **C**. Relative success (y-axis) of progressing from Phase I to Approval for drug-disease pairs with predicted probability above a threshold (x-axis) compared to pairs below the baseline probability (0.004). Values above each point indicate the estimated relative success (fold-change; only when significantly different from 1). Values below points show the number of progressing drug-disease pairs relative to the total evaluated for pairs with predicted probability above the x-axis threshold versus those below the baseline probability. Shaded regions indicate Katz 95% confidence intervals.

We next evaluate the generalizability of our predictions by comparing them with evidence from clinical trials. Although clinical trials data is not used during model training, we find that drug-disease pairs with predicted probabilities higher than the baseline probability of being an indication in the test set (0.004) are more likely to be tested in clinical trials, with the degree of enrichment increasing with predicted probability (**Fig 4B**). When looking at clinical trial progression, we find that drug-disease pairs with higher predicted probabilities are more likely to advance from Phase I to Approval than those with predicted probabilities below the baseline (**Fig 4C**). Here we report relative success (analogous to relative risk) because it captures the proportional difference in trial progression probabilities between groups, which is more interpretable for predicting outcomes conditional on trial initiation. Specifically, pairs with predicted probabilities >0.1 are 2.03 times more likely to progress from Phase I to regulatory approval compared to pairs with probabilities ≤0.004 (**Fig 4C**). While PhaseLI trials test compounds already deemed safe and biologically active in humans, most ultimately fail due to insufficient efficacy; our model is able to distinguish the subset that succeeds. We observe a similar pattern for progression from Phase I to Phase II, Phase II to Phase III, and Phase III to Approval, with the highest relative success being from Phase III to regulatory approval, where tested drugs are compared against standard-of-care treatment (**S8 Fig**).

We provide the full list of drug-disease predicted probabilities of being an indication from both phenotypically dissimilar drug indication-disease pairs, and from all disease pairs in the **Table S1**, along with the corresponding drug indications used for each prediction.

### Integrating multiple measures of genetic similarity in a predictive model for drug side effects

We apply the same approach to estimate the probability that a given drug-disease pair is a side effect. The side effects model using all pairs achieves an AUROC of 0.582 on held-out drug-disease pairs. To evaluate the contribution of genetic similarity as opposed to phenotypic similarity confounding, as previously, we remove drug side effect-disease pairs with phenotypic similarity, re-train our model and re-calculate predicted probabilities. This new model is still predictive of known side effects (AUROC=0.548; individual disease AUROCs shown in **Fig 5A**).

**Fig 5.**
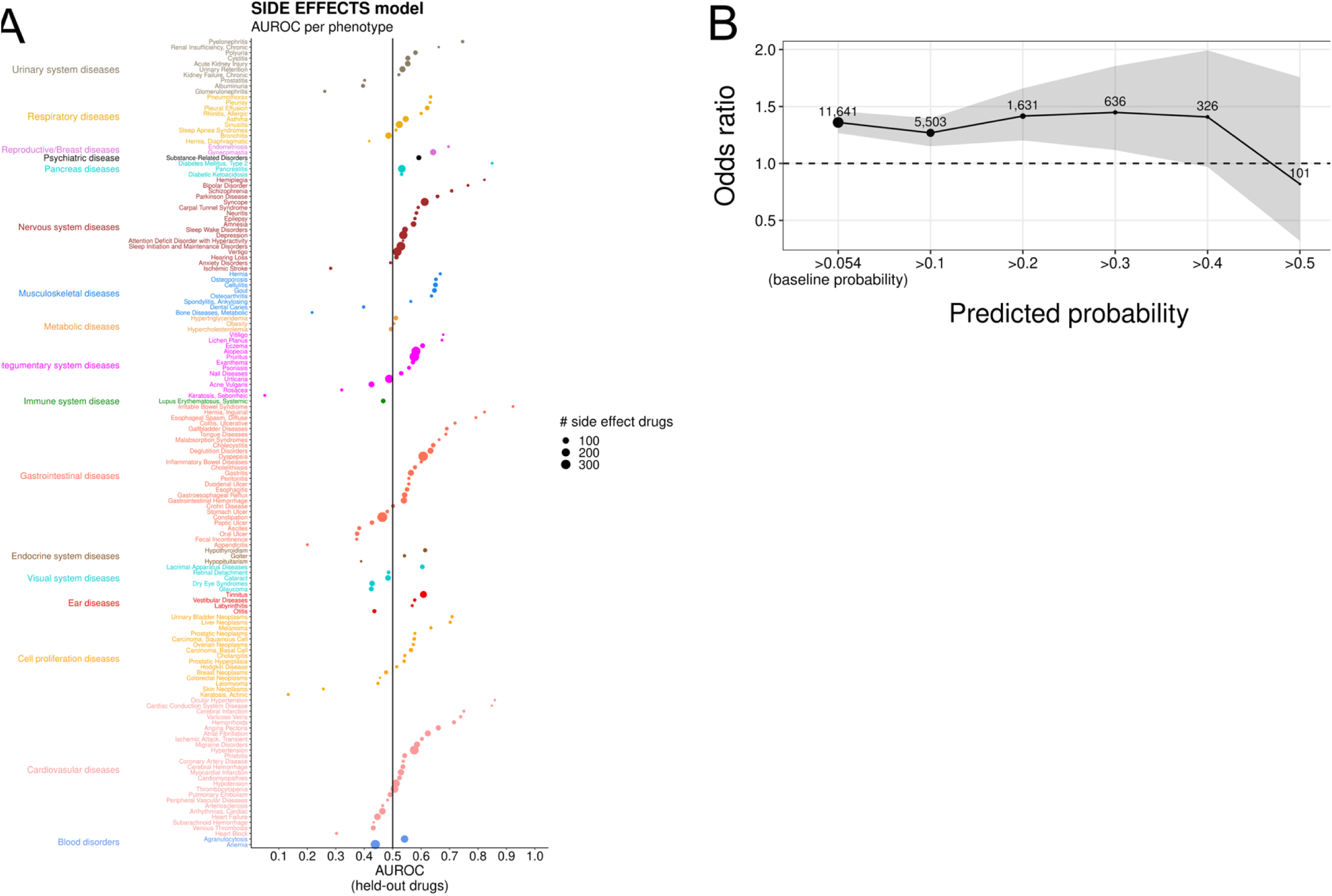
Evaluation of the side effects models. **A.** AUROC (x-axis) per tested disease (y-axis). Color indicates the body system of each disease based on the Open Targets classification. **B.** Odds ratio (y-axis) for drug-disease pairs with predicted probability greater than a given threshold (x-axis) being a side effect (SIDER data) compared to those with predicted probability below the baseline probability of being a side effect in the test set (0.054). Shaded regions indicate 95% confidence intervals calculated by Fisher’s exact test (two-sided). The total number of drug-disease pairs included in this analysis is 43,510 (drugs: 430; diseases: 158). The size of the points is proportional to the number of pairs with predicted probability greater than the x-axis threshold (actual number of pairs is printed above each point).

Next, we assess the generalizability of the side effect predictions derived from the model using phenotypically dissimilar pairs, as our goal is to evaluate the predictive power of genetic similarity. We first use SIDER, a database that extracts drug-side effect relationships from drug labels and black-box warnings(24). After keeping drugs and diseases appearing in both SIDER and onSIDES (our training set) and removing overlapping true labels, we find that pairs with predicted probabilities >0.2 are 1.42 times (95% CI: 1.20-1.66) more likely to be true side effects than those with predicted probabilities below the baseline probability of being a side effect in the test set (0.054). We also compare our predictions to offSIDES, a database that identifies putative drug side effects by mining spontaneous adverse event reports from the FDA Adverse Event Reporting System (FAERS). We observe a similar enrichment, although not significant, potentially due to the low number of predicted drug-disease pairs with offSIDES data available (**S9 Fig**).

We provide the full list of drug-candidate disease predicted probabilities of being a side effect from both approaches (one based on phenotypically dissimilar drug side effect-disease pairs, and one based on all pairs) in the **Table S2**, along with the corresponding drug side effects used for each prediction.

### Genetic similarity points to a shared basis of drug indications and side effects

Finally, we investigate whether indications and side effects of a drug share a common genetic basis. We hypothesize that genetic similarity of a disease to current drug indications can predict whether that disease is a side effect of the drug, and, conversely, similarity to current drug side effects can predict a new indication for that drug. This analysis is motivated by the concept that drugs affect a finite number of biological processes through the genes they target, and these processes may result in either indications or side effects. This concept is also supported by previous studies showing that both drug indications and side effects are enriched for human genetic associations, suggesting a potential genetic link between them(3,4,6). However, this connection remains largely unexplored.

To test this hypothesis, we re-train our indication and side effect models using only drugs and diseases with available information for both. We also exclude drug-disease pairs that are both an indication and a side effect to ensure that any observed signals are not driven by overlapping labels. This results in 667 drugs and 178 diseases (111,474 pairs) available for this analysis. Using these refitted models, we find that the indications model predicts true side effects significantly better than expected by chance (AUROC=0.525, p_permutation_<0.001) (**Fig 6A**). Additionally, we find that the side effects model predicts true drug indications better than random expectations (AUROC=0.542, p_permutation_<0.001) (**Fig 6B**). As side effects are by definition due to a drug’s indication mechanism of action adversely affecting another disease, it is reasonable that side effects could be more informative of indications than the reverse. Notably, since these models only use information about genetic similarity between diseases, these findings indicate that genetic similarity can suggest which drugs have biological effects relevant to the biology of a disease.

**Fig 6.**
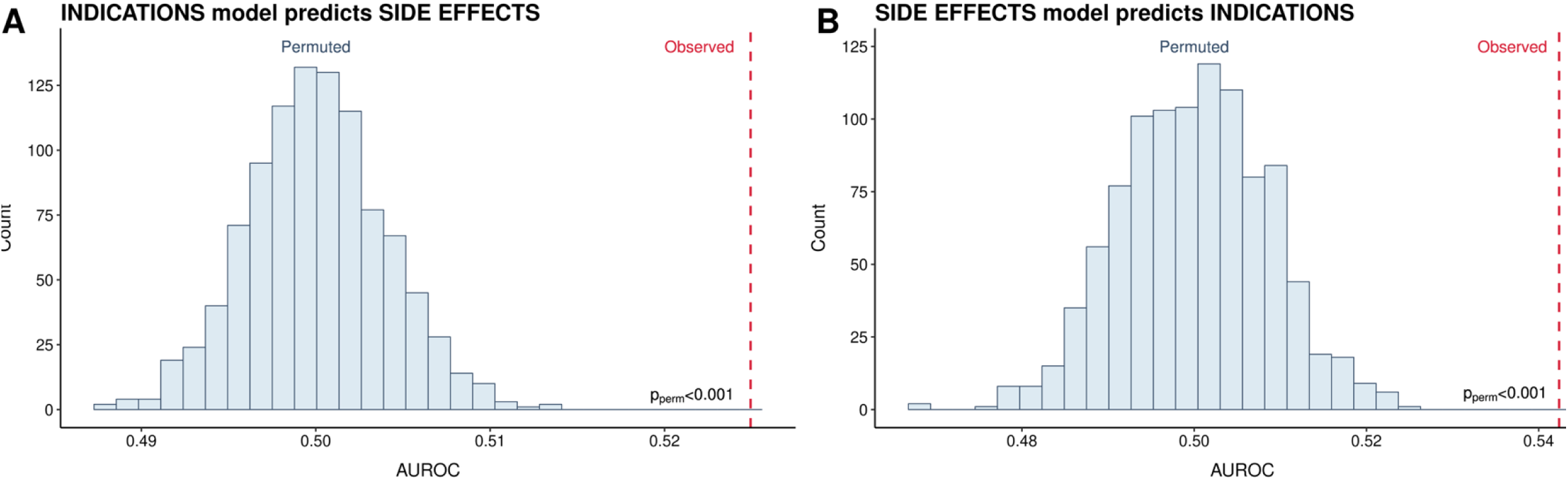
The indication model predicts side effects (A) and vice versa (B). Drug-disease pairs that are both indications and side effects are excluded from model training and evaluation. Models are refitted on common drugs and diseases (total pairs =111,474 pairs; 667 drugs; 178 diseases). Red, vertical dashed lines indicate the observed AUROC in each case. Permutation tests (n=1,000) are conducted by shuffling the true drug-disease labels.

## Discussion

In this work, we develop a genetics-informed framework that leverages pleiotropy to predict new drug indications and side effects. Pleiotropy has previously been used to understand disease etiology (25), and to suggest drug repurposing opportunities among similar diseases. Here, we extend this concept by systematically evaluating whether genetic similarity across a broad range of diseases can uncover drug therapeutic and adverse effects, while accounting for phenotypic similarity confounding. We believe that our work advances the use of genetics for both drug discovery and safety evaluation.

Recent approaches show that GWAS-identified causal genes often represent promising drug targets (3,4). One such approach is the Open Targets L2G machine learning model, which combines different types of data to find the most likely causal gene at a genome-wide significant GWAS locus (p-value<5e-08). Although such approaches have growing support, they are limited because not all drug targets have strong GWAS signals, resulting in missed drug repurposing opportunities. Our method addresses this by enabling predictions even for diseases with weak or missing GWAS signals at drug’s gene-targets. For instance, 29.5% of predicted drug-disease indication pairs with probability p≥0.1 involve drugs whose gene-targets contain at least one subthreshold variant for the disease (GWAS p-value<5e-04), but only 11.3% of our predictions are significant at a genome-wide level (**Supplementary fig S10**). This supports the ability of genetic similarity to identify potential drug-disease indications even when genome-wide significant GWAS hits are absent. Overall, we believe that our approach complements rather than replaces approaches that rely on genome-wide significant GWAS signals.

To illustrate the strength of our approach, we provide an example of a predicted drug-disease pair that is biologically supported but would be overlooked by approaches relying solely on genetic data of the candidate disease. We identify naltrexone as a potential treatment for Irritable Bowel Syndrome (IBS) (predicted probability=0.283) due to its genetic similarity to alcoholism, a naltrexone’s indication. Naltrexone is an antagonist of the μ- (*OPRM1*), κ-(*OPRK1*) and δ- (*OPRD1*) opioid receptors (26). These receptors, besides in the brain, are also expressed in the gastrointestinal tract, where they play a role in regulating motility, secretion, and visceral sensation, all processes believed to be implicated in IBS pathophysiology (27). This biological rationale supports our prediction. Additionally, there is evidence from prior studies, including one pilot clinical study, suggesting that naltrexone may treat IBS by reducing neuroinflammation and visceral hypersensitivity (28–30). Notably, this repurposing candidate would have been missed by approaches looking solely at the IBS GWAS since there is currently no genetic evidence linking IBS to any of naltrexone known gene-targets (*OPRM1*, *OPRK1*, *OPRD1*; based on data from the Open Targets Platform v25.09, https://platform.opentargets.org/).

A similar insight applies to side effects. We predict that anastrozole may cause malabsorption syndrome (predicted probability=0.61) based on its genetic similarity to anastrozole’s known side effects, osteoporosis and phlebitis. Anastrozole is an aromatase inhibitor that blocks *CYP19A* and thereby prevents the conversion of androgens to estrogens (31). This reduces the systemic estrogen levels which explains anastrozole’s established adverse effects on bone and vascular health (32–34). Notably, estrogen receptors are also expressed in the gastrointestinal tract, where they regulate nutrient absorption, including calcium and vitamin D (35–37). Consequently, suppression of estrogen signaling by anastrozole could impair intestinal absorption. Together, these reflect a shared genetic link between osteoporosis, phlebitis and malabsorption syndrome tied to estrogen signaling, highlighting the ability of our genetics-informed side effect model to uncover mechanistically coherent drug-disease relationships across diverse clinical contexts.

We also find evidence for a shared genetic basis between drug indications and side effects, as our indication model can predict side effects, and vice versa, better than expected by chance. This suggests that a drug’s therapeutic benefits and side effects may be due to the drug’s effect on biological pathways that influence multiple disease phenotypes. For instance, our side effect model predicts mercaptopurine, a purine antagonist, to have an effect on non-Hodgkin lymphoma (predicted probability=0.52) based on its genetic similarity to mercaptopurine’s known side effects, such as anemia and alopecia. Mercaptopurine impairs DNA replication and induces apoptosis in rapidly dividing cells by inhibiting purine synthesis (38). While this mechanism of action underlies its known toxicities (anemia due to suppression of hematopoietic progenitors; alopecia due to inhibition of hair follicle keratinocytes) it also suggests efficacy against other highly proliferative cell populations, such as malignant lymphocytes in non-Hodgkin lymphoma. Therefore, this prediction reflects a cross-system biological pleiotropy involving a shared proliferative and purine-dependent axis, in which modulation of the same molecular pathway confers side effects in normal tissues while providing therapeutic benefit in a proliferative malignancy.

Our study has several limitations. First, while we use five complementary metrics to define genetic similarity and predict novel drug therapeutic and adverse effects, many drugs treat disease symptoms rather than underlying biological causes. Such symptomatic treatments may not be captured by genetic associations and could therefore be missed by our approach. Second, the incompleteness of publicly available genetic datasets limits our ability to compute genetic similarity scores for all drug-candidate disease pairs, thereby preventing a fair comparison of the relative predictive power of each developed metrics. Although imputation could address this limitation, we chose not to pursue it due to variation in the extent and nature of missingness across datasets. Third, the GWAS sample size varies across diseases and even for the same disease between different genetic databases. For example, GWAS Atlas and PhenomeXcan may use different GWASs for the same disease. Diseases with smaller GWAS may yield less confident similarity estimates, which could potentially cause us to overlook some drug-disease links. Fourth, our developed measures do not capture all genetic similarity between disease pairs. Some of our measures rely on significant genes or loci, which, as mentioned above, must be conservative and can miss some signal. Genome-wide measures like LD-score correlation, conversely, can miss more localized similarity. Future work could identify a wider range of measures of similarity. Fifth, there is a confounding association between genetic similarity and phenotypic similarity, and the latter is already commonly exploited to repurpose drugs for physiologically similar diseases. To isolate the utility of genetic similarity, we stringently remove pairs of diseases with obvious clinical similarity using both disease annotations and knowledge graph similarities. This also addresses potential sample overlap between GWAS as phenotypically similar diseases tend to be clinically associated. As a result, we are able to show that genetic similarity enriches for uses of drugs in phenotypically dissimilar diseases. We also supply the full results in the supplement. Therefore, we believe that this limitation does not invalidate the identified associations.

In conclusion, our work emphasizes the potential of genetic similarity to inform drug discovery and overcome known limitations of single disease GWAS studies. To our knowledge, this is the first study that evaluates the use of genetic similarity to inform drug discovery at a phenome-wide scale. Future work could use our predictions as a resource for experimental validation, as well as could use complete genetic data to fairly compare the predictive performance of individual genetic similarity metrics and reveal which data sources are most informative for making drug-disease predictions. Furthermore, future studies can build on our findings of shared genetic basis between drug indications and side effects to identify biological pathways that drive both drug efficacy and toxicity. Ultimately, such insights could help reduce clinical trial attrition rates by prioritizing candidate drugs with a more favorable side effect profile.

## Materials and Methods

### Genetic data and development of genetic similarity metrics

We define genetic similarity between two diseases as the extent to which they share genetic variants or genes. To quantify this, we develop five genetic similarity metrics, each derived from a different type of genetic data. Below, we describe each metric and its data source.

1. <u>LDSC-based metric:</u> our first metric captures genome-wide genetic correlation between disease pairs using LD Score Regression (LDSC) (25). We obtain precomputed LDSC genetic correlation estimates from the GWAS Atlas (https://atlas.ctglab.nl/; release 3, v20191115; gwasATLAS_v20191115_GC.txt.gz), covering 2,415 pairs across 70 diseases(10). These values are used as is without further processing.
2. <u>MAGMA-based metric:</u> our second metric uses output statistics from MAGMA, a tool that aggregates SNP-level GWAS signals within genes to generate gene-level p-values for association with a disease (MAGMA gene analysis) (39). We obtain precomputed MAGMA results from the GWAS Atlas (https://atlas.ctglab.nl/; release 3, v20191115; gwasATLAS_v20191115_magma_P.txt.gz) for 81 diseases and 18,680 genes(10). However, this file includes missing values for some disease-gene pairs. To address this without losing the majority of information, we exclude the top-10 diseases with the most missing values (>10%) and keep genes with complete data for all the remaining diseases. This results in a dataset of 71 diseases and 15,297 genes. To retain informative genes, we keep only those significantly associated with at least one disease (BH-adjusted p<0.05), yielding 10,302 genes. Then, for each disease pair, we calculate the spearman correlation between their gene-level association p-values.
3. <u>S-MultiXcan-based metric:</u> our third metric is based on S-MultiXcan, a tool that summarizes genetically predicted gene expression across 44 GTEx tissues to estimate gene-disease associations (40). We obtain precomputed S-MultiXcan p-values from PhenomeXcan (https://zenodo.org/records/3911190; smultixcan-mashr-pvalues.tsv.gz), covering 137 diseases and 22,215 genes. We convert p-values into z-scores, as described elsewhere (41). Then, for each disease pair, we compute the spearman correlation between their gene regulation profiles.
4. <u>L2G-based metric:</u> our fourth metric uses data from the Open Targets Locus-to-Gene (L2G) model (23). L2G is a machine learning model that assigns a probability to each gene to be causal for a disease based on variants in a genome-wide significant GWAS locus and evidence from gene proximity, QTL colocalization, chromatin interactions, and variant pathogenicity. We download L2G scores for 137 diseases and 14,951 genes from the Open Targets Genetics platform (v22.09; https://ftp.ebi.ac.uk/pub/databases/opentargets/genetics/22.09/l2g/). In case of duplicated disease-gene entries, we keep the one with the highest L2G score. For disease-gene pairs with no available L2G score, we assign a value of 0, reflecting no evidence of causality. Finally, for each disease pair, we compute the spearman correlation between their L2G gene-level profiles.
5. <u>COLOC-based metric:</u> our fifth and final metric uses colocalization data between GWAS loci and molecular QTLs (eQTLs, pQTLs, sQTLs). We obtain this data from the Open Targets Genetics platform (v22.09; https://ftp.ebi.ac.uk/pub/databases/opentargets/genetics/22.09/v2d_coloc/). For each disease, we identify genes with strong colocalization support (posterior probability for shared causal variant, H4 ≥ 0.8, coloc), yielding colocalization-based gene sets for 39 diseases (total of 1,758 genes). Then, for each disease pair, we quantify genetic similarity using the overlap coefficient, defined as:

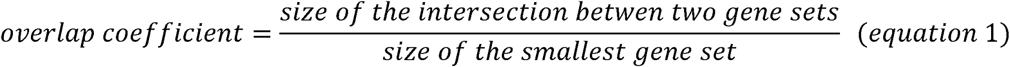

To ensure consistent disease mapping across all five genetic similarity metrics, we perform manual curation of disease identifiers. For each of the 178 diseases, we retrieve the corresponding MeSH ID and then match it to the relevant identifier in each genetic data source. In cases where multiple GWAS entries correspond to the same disease, we select the most recent GWAS or the one with the largest sample size.

Each data source quantifies association of diseases with a variable portion of the genome. Open Targets computes L2G scores only for genome-wide significant GWAS loci for a disease and COLOC analysis is performed only when these loci overlap with known molecular QLTs. If no such overlap exists, the locus is excluded from COLOC analysis.

Due to differences in data availability across sources, coverage varies by disease. Nevertheless, the vast majority of diseases (173/178; 97.2%) are represented in at least two of the three sources. Within each source, coverage also varies across data types which is a result of source-specific data processing pipelines (explained in the documentation of each resource). For example, within Open Targets, we have L2G data for 137 diseases, but we have COLOC data for 51 of them, depending on the number of GWAS loci passing genome-wide significance.

### Disease annotations and phenotypic similarity data

We make extensive efforts to disentangle genetic similarity from clinical similarity in order to assess the additive value of genetic similarity for drug discovery. To this end, we compile disease annotations from Open Targets and complement them with manually curated labels from ICD10 and the UK HRCS. For ICD10, we map each disease to its broadest diagnostic category. For example, we assign type 2 diabetes (ICD10 code: E11) to the E00-E89 category, corresponding to “Endocrine, nutritional and metabolic diseases”. For UK HRCS, we manually assign each disease to the most relevant health category based on online guidelines (https://hrcsonline.net/health-categories/). Using these annotations, we classify each disease pair as affecting the same body system if the two diseases share a category in at least one of the annotation sources or affecting different body systems otherwise.

Diseases affecting different body systems can still be phenotypically similar. For instance, hypercholesterolemia (metabolic disease) and myocardial infarction (cardiovascular disease) are classified as affecting different body systems using standard disease annotations, yet they have a well-established causal relationship. Therefore, we also use ClinGraph, a large-scale clinical knowledge graph that integrates eight standardized medical vocabularies to represent relationships among diseases, symptoms, drugs, and clinical concepts. We use pre-trained disease embeddings from ClinGraph (https://zitniklab.hms.harvard.edu/projects/Clinical-knowledge-embeddings/) and quantify phenotypic similarity between disease pairs as the cosine similarity of these embeddings. For diseases that map to multiple ClinGraph identifiers, we compute their phenotypic similarity to another disease as the mean cosine similarity across all corresponding identifier pairs. Supplementary figure 3 shows the distribution of ClinGraph phenotypic similarity values across disease pairs affecting same or different body systems according to standard disease annotations.

### Drug indications data

We compile a comprehensive dataset of approved drug indications by combining data from three publicly available resources: ChEMBL, RxNORM, and SIDER.

1. <u>ChEMBL:</u> we download drug-disease indication data using the ChEMBL API (https://www.ebi.ac.uk/chembl/api/data/drug_indication.json) and filter for drug-disease pairs with “max_phase_for_ind” equal to 4 or labeled as “Approved”. This ensures that only already approved indications are included.
2. <u>RxNORM:</u> we first retrieve a list of drugs mapping RxNORM names to ChEMBL IDs from the UniChem EBI (https://ftp.ebi.ac.uk/pub/databases/chembl/UniChem/data/wholeSourceMapping/src_id1/src1src47.txt.gz). Using the RxNORM names of these drugs, we query the RxNORM API (byDrugName function) to obtain diseases labeled as “may_treat” or “may_prevent” for each drug. Finally, we map the disease IDs to MeSH IDs using the UMLS API for consistency across data sources.
3. <u>SIDER:</u> we download drug indication data from the SIDER database (http://sideeffects.embl.de/media/download/meddra_all_indications.tsv.gz). In SIDER, drug IDs are in PubChem and disease IDs are UMLS CUIs. We convert drug IDs to ChEMBL IDs using the corresponding UniChem mapping file (https://ftp.ebi.ac.uk/pub/databases/chembl/UniChem/data/wholeSourceMapping/src_id1/src1src5.txt.gz) and disease IDs to MeSH IDs using the UMLS API.

We combine drug indication data across all three resources, remove duplicate entries, and exclude overly broad disease categories (top MeSH headers), such as “cardiovascular diseases” and “nervous system diseases”.

### Drug side effects data from drug labels

We obtain drug side effects data from the onSIDES (v3.1.0; https://github.com/tatonetti-lab/onsides/releases/download/v3.1.0/onsides-v3.1.0.zip), a database that extracts adverse events from drug labels using natural language processing. We join the “product_adverse_effect.csv” and “product_label.csv” files to create a table with drug-side effects, and filter for those predicted to be side effects (pred1>pred0). Then, we use UniChem EBI to convert drug RxNORM names to ChEMBL IDs (https://ftp.ebi.ac.uk/pub/databases/chembl/UniChem/data/wholeSourceMapping/src_id1/src1src47.txt.gz) and the UMLS API (crosswalk function) to convert disease MedDRA IDs to MeSH IDs.

We also obtain drug side effects data from SIDER (v4.1; http://sideeffects.embl.de/). Using a similar approach, we map drug PubChem IDs to ChEMBL IDs (UniChem EBI) and disease UML CUIs to MeSH CUIs (UMLS API). We use these drug-disease side effects as an independent dataset to evaluate our side effects model predictions, after removing overlapping labels with onSIDES.

### Drug overlap metrics among diseases

To quantify the overlap of indicated or side effect drugs among diseases, we use the aforementioned compiled drug datasets, and, for each disease pair, we estimate the overlap coefficient of the drugs that treat them or cause them as a side effect (**equation 1**).

### Drug-disease pairs in clinical trials

To find which of the analyzed drug-candidate disease pairs in our sample are currently being, or have been, tested in clinical trials, we download clinical trial data from ChEMBL and the Aggregate Content of https://ClinicalTrials.gov (AACT) database. AACT is a publicly available relational database that contains information about all the trials registered in https://ClinicalTrials.gov (42). We obtain information for all clinical trials that were registered in https://ClinicalTrials.gov as of November 4, 2022. For each drug-disease pair, we keep the maximum clinical trial phase reached. We group clinical trial phases to Phase I (Phase I and Early Phase I), Phase II (Phase II and Phase I/Phase II), Phase III (Phase III and Phase II/Phase III) or unknown phase (no phase information provided). Clinical trials in Phase 0 are at the pre-clinical stage and therefore excluded. Then, we map all drug MeSH IDs to ChEMBL IDs through DrugBank IDs using the UMLS API (crosswalk function) and a mapping file provided by UniChem EBI (https://ftp.ebi.ac.uk/pub/databases/chembl/UniChem/data/wholeSourceMapping/src_id1/src1src2.txt.gz). We also map all disease IDs to MeSH IDs using the UMLS API. We use this data to evaluate our indication model predictions.

### Drug side effect data from spontaneous reports

To evaluate our side effect model predictions, we use data from offSIDES (http://github.com/tatonetti-lab/offsides/), a resource that identifies putative drug-side effect associations by detecting disproportionality signals in spontaneous adverse event reports from FAERS while adjusting for potential confounders (43). For each drug-disease pair, offSIDES reports a Proportional Reporting Ratio (PRR) and an associated p-value, quantifying whether a side effect is reported more frequently for a given drug compared with a matched background of reports for other drugs. We filter the offSIDES data for drug-side effect pairs with PRR>1, indicating an increased reporting frequency for the tested drug. We further filter for drugs and side effects present in our study. Drug-disease pairs with a Bonferroni adjusted p-value<0.05 are considered potentially true side effects. Finally, we convert drug PubChem IDs to ChEMBL IDs using the PubChem Exchange Identifier and disease UMLS CUIs to MeSH IDs using the UMLS API. This yields 20,057 pairs of 362 drugs and 159 diseases, of which 11,505 pairs are identified as significant side effects (Bonferroni p.adjust <0.05).

### Bayesian logistic regression model incorporating all genetic similarity data

To integrate our measures of genetic similarity in a model to predict the probability that a given drug-disease pair represents a potential therapeutic indication, we train a Bayesian logistic regression model using Stan (rstan v2.32.7). We choose this modeling approach because: 1) its probabilistic framework allows the number of disease-indication pairs with genetic similarity to vary across genetic evidence, drugs, and diseases and 2) it incorporates all genetic similarity metrics along with the similarity of a disease to all indications of a drug, rather than only the most genetically similar drug indication.

Our model predicts for a disease i, drug j pair (Y_ij_), the probability p_ij_ of the drug treating the disease. The prediction is modeled using the set of genetic similarities for each evidence type *e* available, assessing similarity of disease *i* to the N annotated indications {*g_ije1_, …, g_ijeN_*}. The genetic similarity scores are approximately normally distributed, and the known drug-treatment labels are of course binary. Therefore, we model:

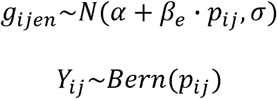

The learned parameters are,e,pij, while is treated as a hyperparameter. To find the best σ, we test several values by training the model on a set of drug-indication pairs and evaluating its performance on held-out drug-indication labels, in a cross validation-like setup. Ultimately, only the predicted probabilities for held-out drug-disease pairs are used for evaluating our model. The full Stan model code is available on our GitHub repository (https://github.com/lalagkaspn/genetic_similarity_drug_discovery).

We train one Bayesian logistic regression model using selected hyperparameters and genetic similarity data across all drug indication-disease pairs (indications model). To account for confounding by phenotypic similarity between diseases and isolate the signal from genetic similarity, we also train a second model using the same hyperparameters but restricted to phenotypically dissimilar drug indication-disease pairs. We define a pair of diseases as phenotypically dissimilar if they do not affect the same body system in any disease annotation and have ClinGraph phenotypic similarity <-0.1 (see “Disease annotations and phenotypic similarity data” for more details). We follow the same process to train the side effect model.

### Permutation tests for the cross-model prediction analysis

To evaluate whether the observed associations in our cross-prediction analysis are due to chance, we perform permutation tests. Specifically, we randomly shuffle the true drug-disease labels (indications or side effects) to generate new pairings that disrupt any real associations and calculate the AUROC. We repeat this process 1,000 times to estimate a null distribution of AUROCs. We then compare the observed AUROC to the null distribution and calculate a permutation p-value as the proportion of permuted AUROCs that are greater than or equal to the observed AUROC. Observed associations are considered statistically significant if the permutation p-value is less than 0.05.

## Declarations

### Ethics approval and consent to participate

Not applicable

### Consent for publication

Not applicable

### Data availability

Data and code for reproducing the results and figures presented in this study can be found in this GitHub repository: https://github.com/lalagkaspn/genetic_similarity_drug_discovery. All drug-disease predictions are included within the article and its Supplementary Tables S1 and S2.

### Completing interests

Not applicable

### Authors’ contributions

PNL curated and analyzed the genetic and drug data. RDM conceptualized and supervised the research. PNL and RDM interpreted the results and, drafted and revised the manuscript. All authors read and approved the final manuscript.

## Acknowledgements

Not applicable

