## Supplementary Figures for "Genetic similarity among 178 disease phenotypes predicts therapeutic and side effects for 1,711 drugs"

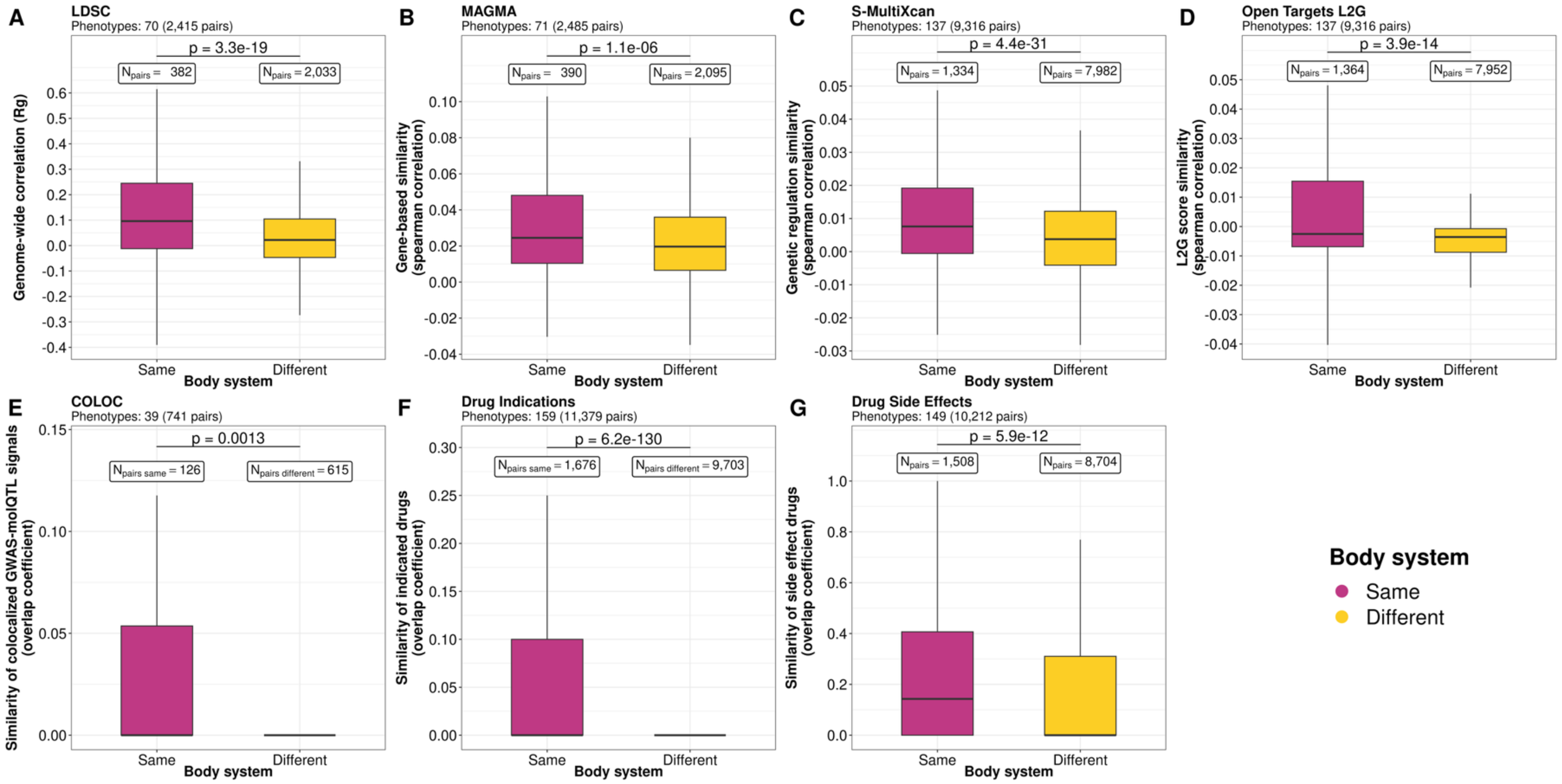


**Supplementary fig S1. Developed genetic similarity and drug overlap metrics capture known biology. A-E.** Disease pairs affecting the same body system have higher genetic similarity scores than those affecting different body systems, across five genetic similarity metrics: LDSC (A), MAGMA (B), S-MultiXcan (C), L2G (D), and COLOC (E). **F-G**. Disease pairs within the same body system share more indicated (F) and side effect (G) drugs than those from different body systems. We derive body system annotations from Open Targets, ICD-10, and UK HRCS. We classify a disease pair as affecting the same body system if both diseases are assigned to a common system in any of these databases.

In all panels, statistical comparisons between groups are performed using the Wilcoxon rank-sum test (two-sided). Due to the incompleteness of publicly available sources used, the number of diseases included in each analysis differs and is mentioned in the title of each plot. In all panels, outliers, defined as values below Q1−1.5×IQR or above Q3+1.5×IQR, are not shown but are included in the statistical analyses.

Abbreviations: LDSC, Linkage Disequilibrium Score Regression; MAGMA, Multi-marker Analysis of GenoMic Annotation; L2G, Locus-to-Gene; Q1: 25th percentile; Q3: 75th percentile; IQR: interquartile range; UK HRCS: United Kingdom Health Research Classification System; ICD-10: International Classification of Diseases-Tenth version

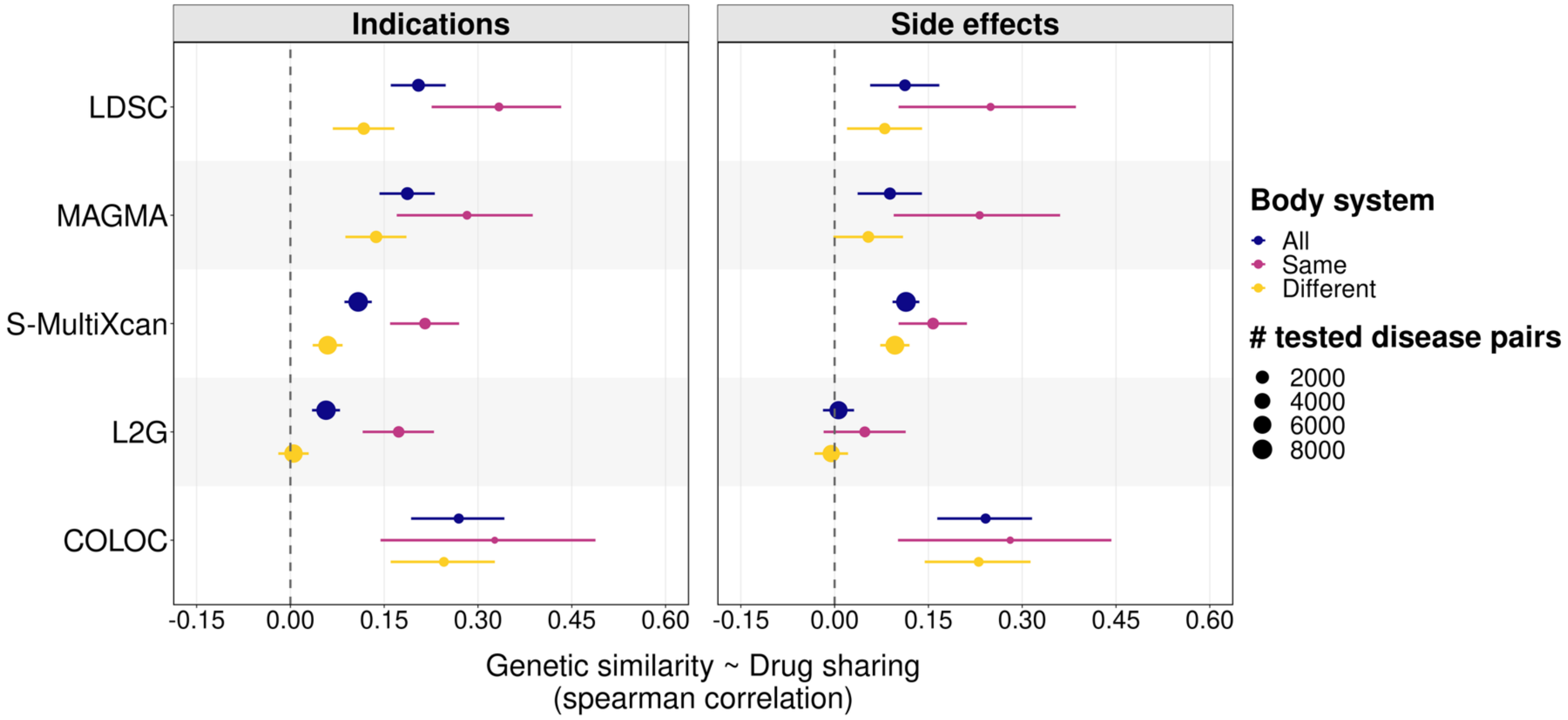


**Supplementary fig S2. Diseases with higher genetic similarity scores share more drugs, either as therapeutic indications or side effects, even after stratification by body system.** Each point represents the spearman correlation between a genetic similarity metric (y-axis) and drug sharing across disease pairs (left panel: sharing of drug indications; right row: sharing of drug side effects). The size of the points is proportional to the number of disease pairs contributing in each analysis (color). Disease annotations are obtained from Open Targets, ICD-10 and UK HRCS.

Abbreviations: UK HRCS: United Kingdom Health Research Classification System; ICD-10: International Classification of Diseases-tenth version

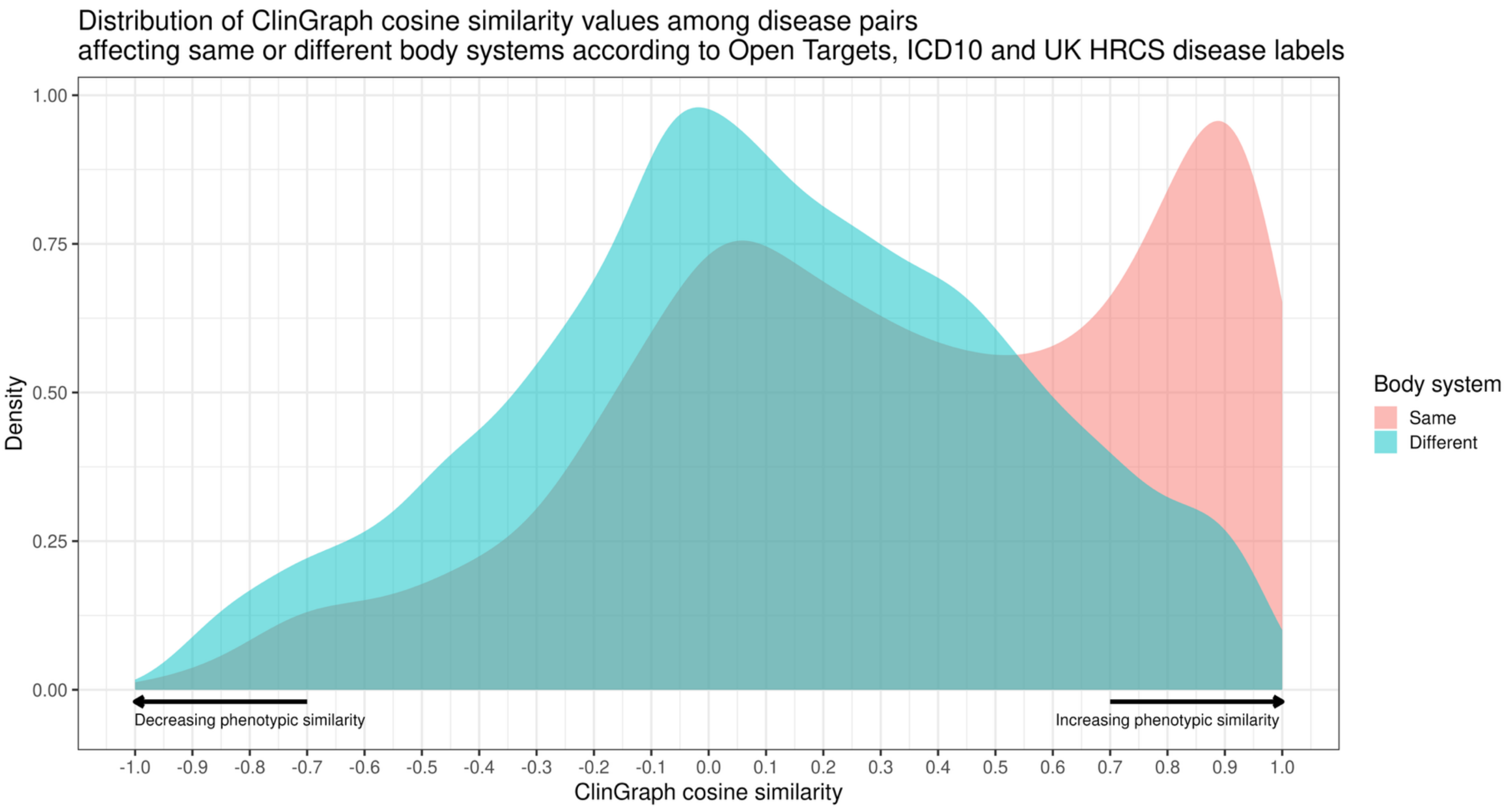


**Supplementary fig S3. Distribution of ClinGraph embedding cosine similarity values (phenotypic similarity) for disease pairs affecting the same versus different body systems**. As expected, disease pairs affecting the same body systems have higher ClinGraph phenotypic similarity than those affecting different body systems. We derive body system annotations from Open Targets, ICD-10, and UK HRCS. We classify a disease pair as affecting the same body system if both diseases are assigned to at least one common system in any of these databases.

Abbreviations: UK HRCS: United Kingdom Health Research Classification System; ICD-10: International Classification of Diseases-tenth version

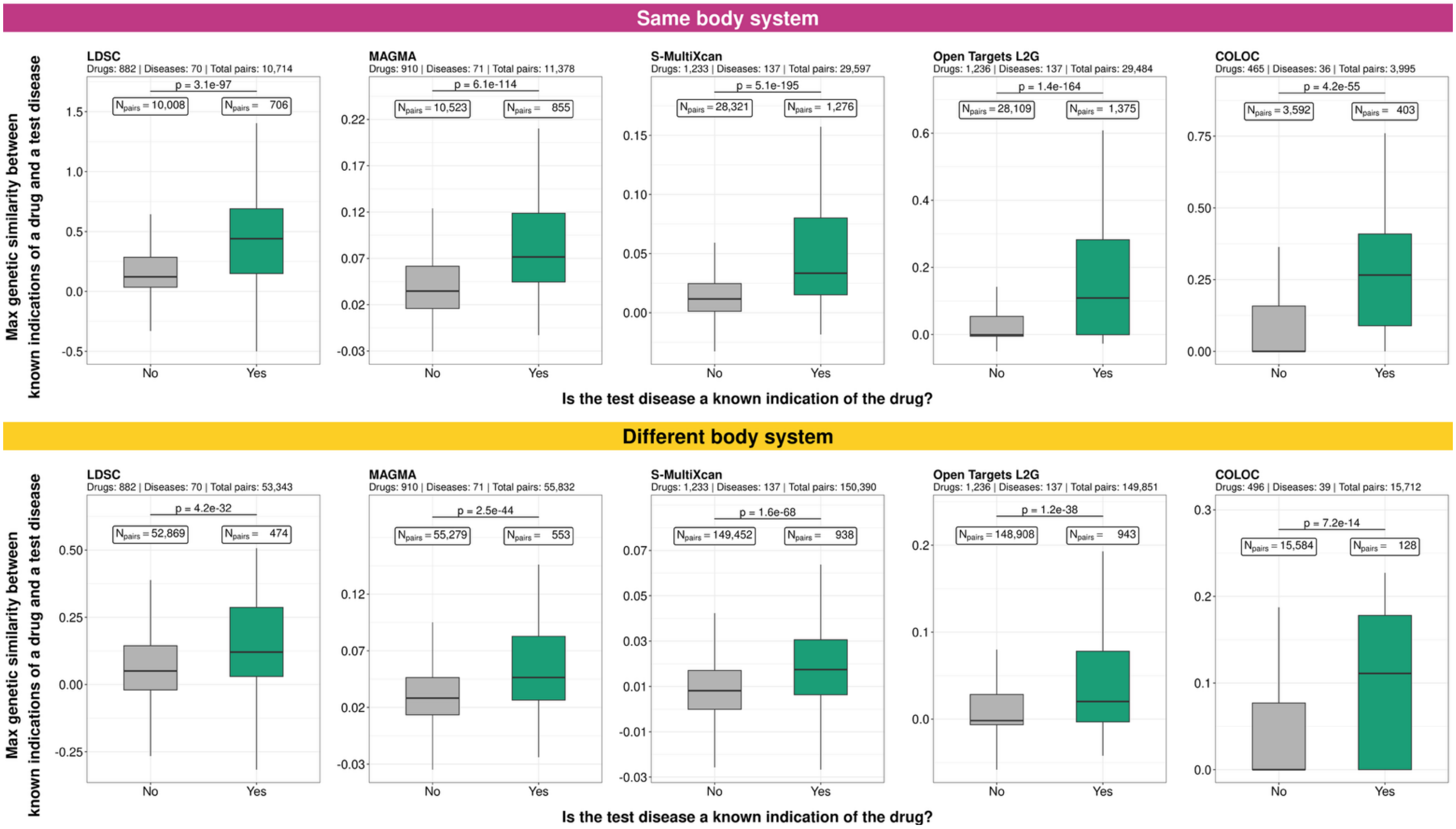


**Supplementary fig S4. Genetic similarity distinguishes known drug indications even after stratification by body system.** Y-axis: maximum genetic similarity between a drug's known indications and a test disease. Top-panel: subset of drug known indications-disease pairs affecting the same body system. Bottom-panel: subset of drug known indications-disease pairs affecting different body systems. We derive disease body system annotations from Open Targets, ICD-10, and UK HRCS. We classify a drug indication-disease pair as affecting the same body system if both are assigned to at least one common system in any of these databases. Color denotes whether the drug-disease pair is a known indication (green) or not (gray). Outliers, estimated as values below Q1 − 1.5×IQR or above Q3 + 1.5×IQR, are not shown but included in the statistical analysis (Wilcoxon rank-sum test, two-sided).
IQR: Inter-quartile range; Q1: first quartile (25th percentile); Q3: third quartile (75th percentile)
Abbreviations: UK HRCS: United Kingdom Health Research Classification System; ICD-10: International Classification of Diseases-tenth version

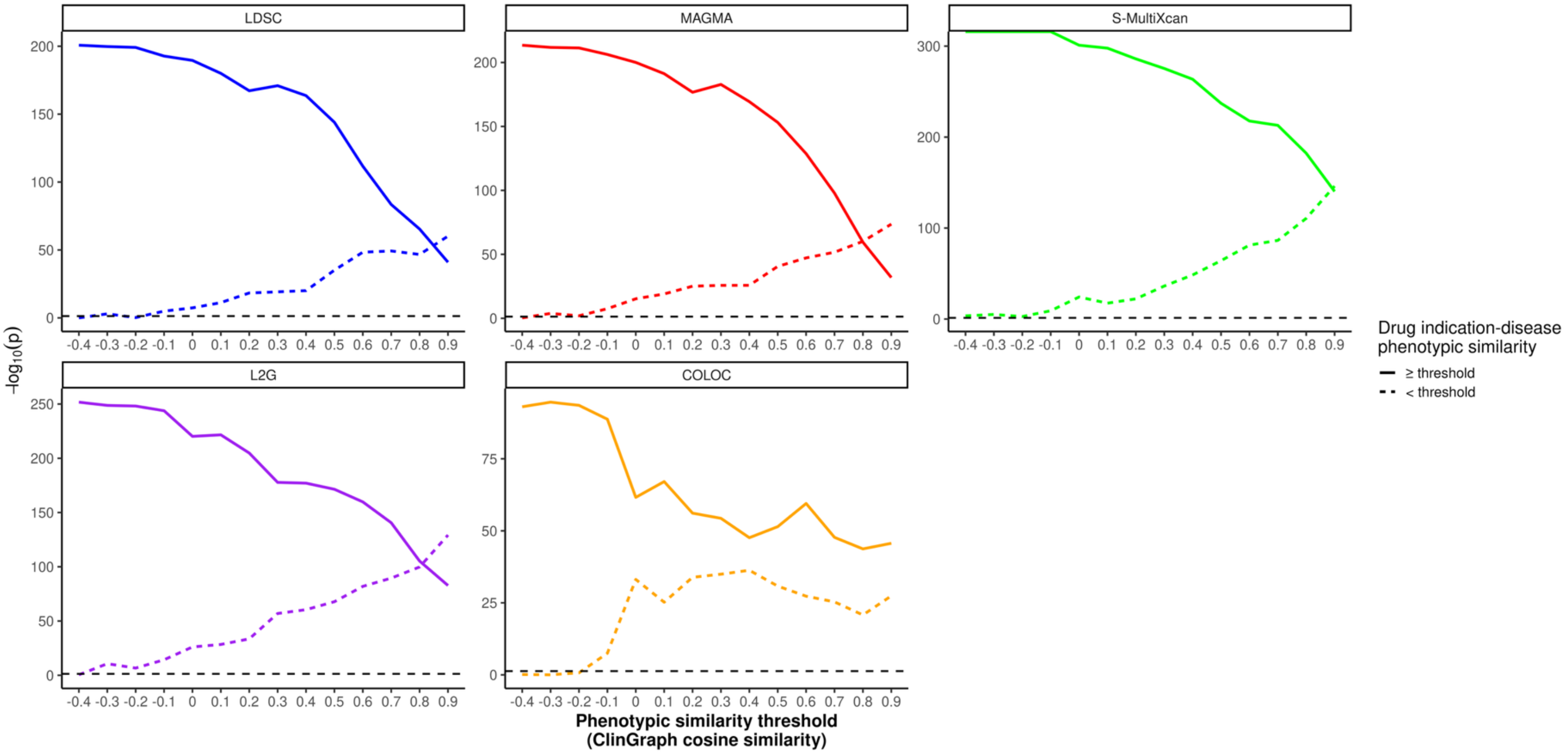


**Supplementary fig S5. Genetic similarity distinguishes known drug indications even after stratification by phenotypic similarity.** Y-axis: -log10(p-value) of Wilcoxon rank-sum test (one-sided) testing whether diseases treated by the same drugs are more genetically similar than diseases that do not. The solid line represents the analysis where only drug indication-disease pairs with phenotypic similarity (ClinGraph embedding cosine similarity) greater than or equal to a x-axis threshold are included. The dashed line is from the analysis where pairs with cosine similarity below a x-axis threshold. Color denotes the genetic similarity metric used.

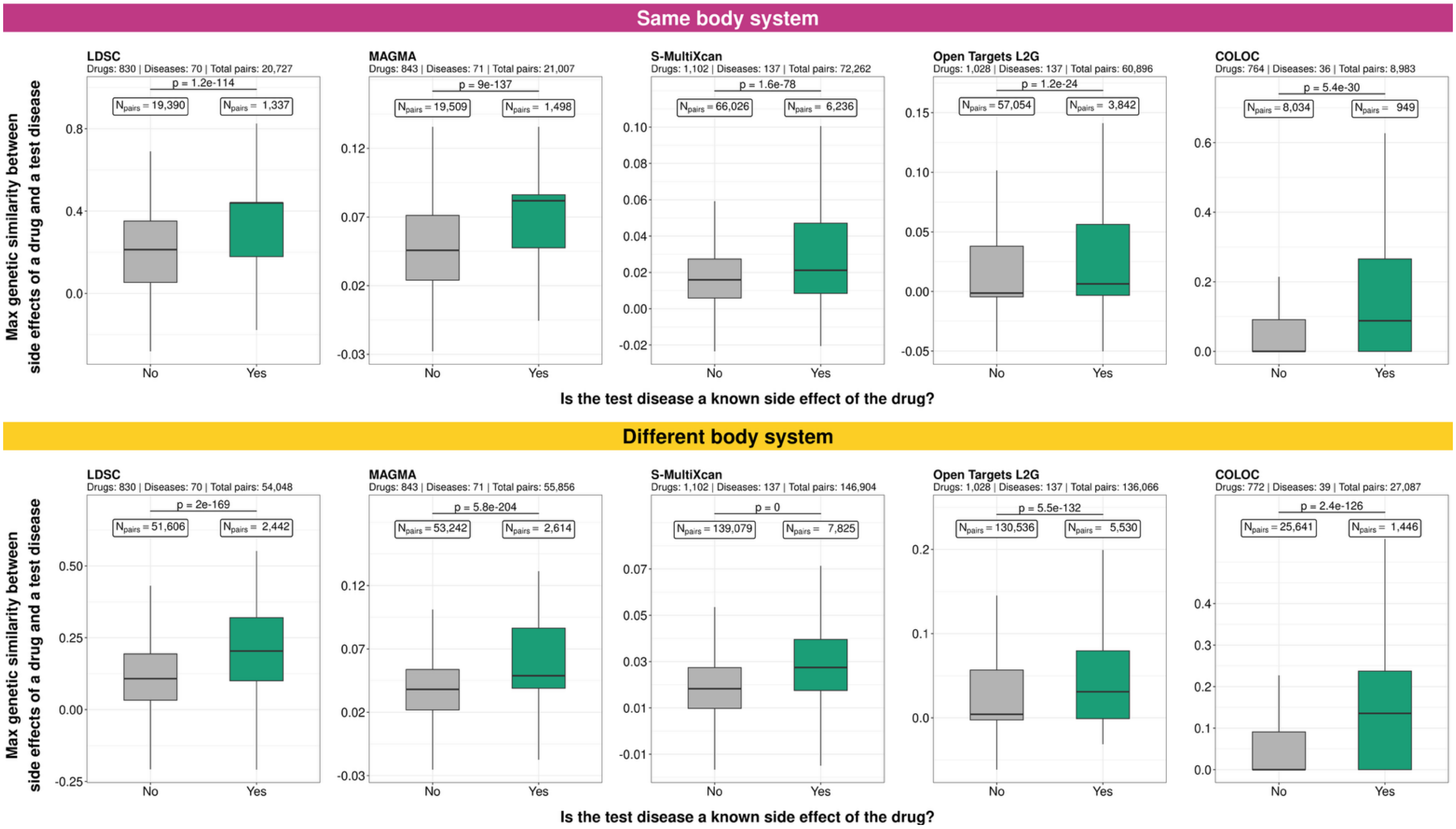


**Supplementary fig S6. Genetic similarity distinguishes known drug side effects even after stratification by body system.** Y-axis: maximum genetic similarity between a drug's known side effects and a test disease. Top-panel: subset of drug known side effect-disease pairs affecting the same body system. Bottom-panel: subset of drug known side effect-disease pairs affecting different body systems. We derive disease body system annotations from Open Targets, ICD-10, and UK HRCS. We classify a drug side effect-disease pair as affecting the same body system if both are assigned to at least one common system in any of these databases. Color denotes whether the drug-disease pair is a known side effect (green) or not (gray). Outliers, estimated as values below Q1 − 1.5×IQR or above Q3 + 1.5×IQR, are not shown but included in the statistical analysis (Wilcoxon rank-sum test, two-sided).
IQR: Inter-quartile range; Q1: first quartile (25th percentile); Q3: third quartile (75th percentile)
Abbreviations: UK HRCS: United Kingdom Health Research Classification System; ICD-10: International Classification of Diseases-tenth version

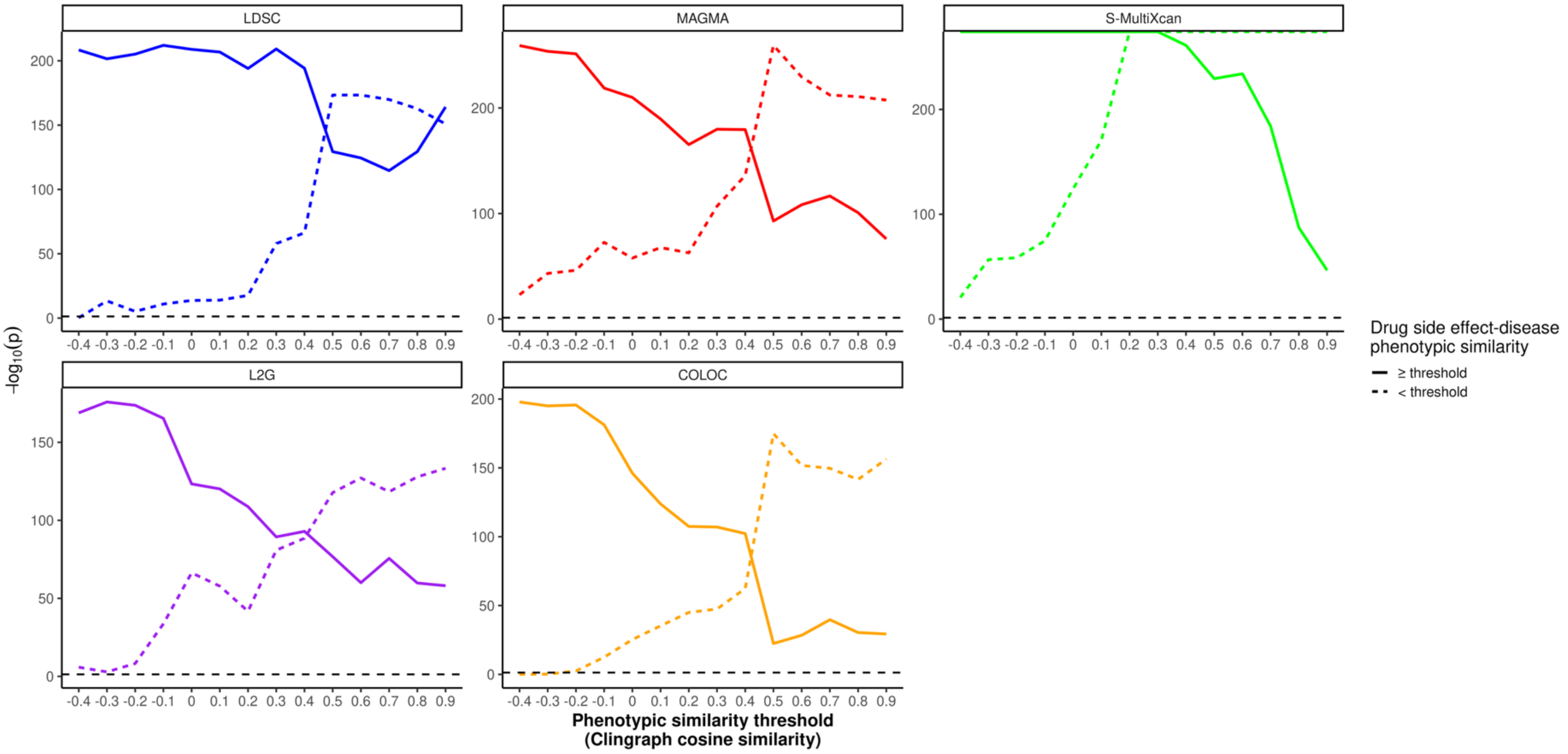


**Supplementary fig S7. Genetic similarity distinguishes known drug side effects even after stratification by phenotypic similarity.** Y-axis: -log10(p-value) of Wilcoxon rank-sum test (one-sided) testing whether diseases caused as side effects by the same drugs are more genetically similar than diseases that do not. The solid line represents the analysis where only drug side effect - disease pairs with ClinGraph embedding cosine similarity greater than or equal to a x-axis threshold are included. The dashed line is from the analysis where pairs with cosine similarity below a x-axis threshold. Color denotes the genetic similarity metric used.

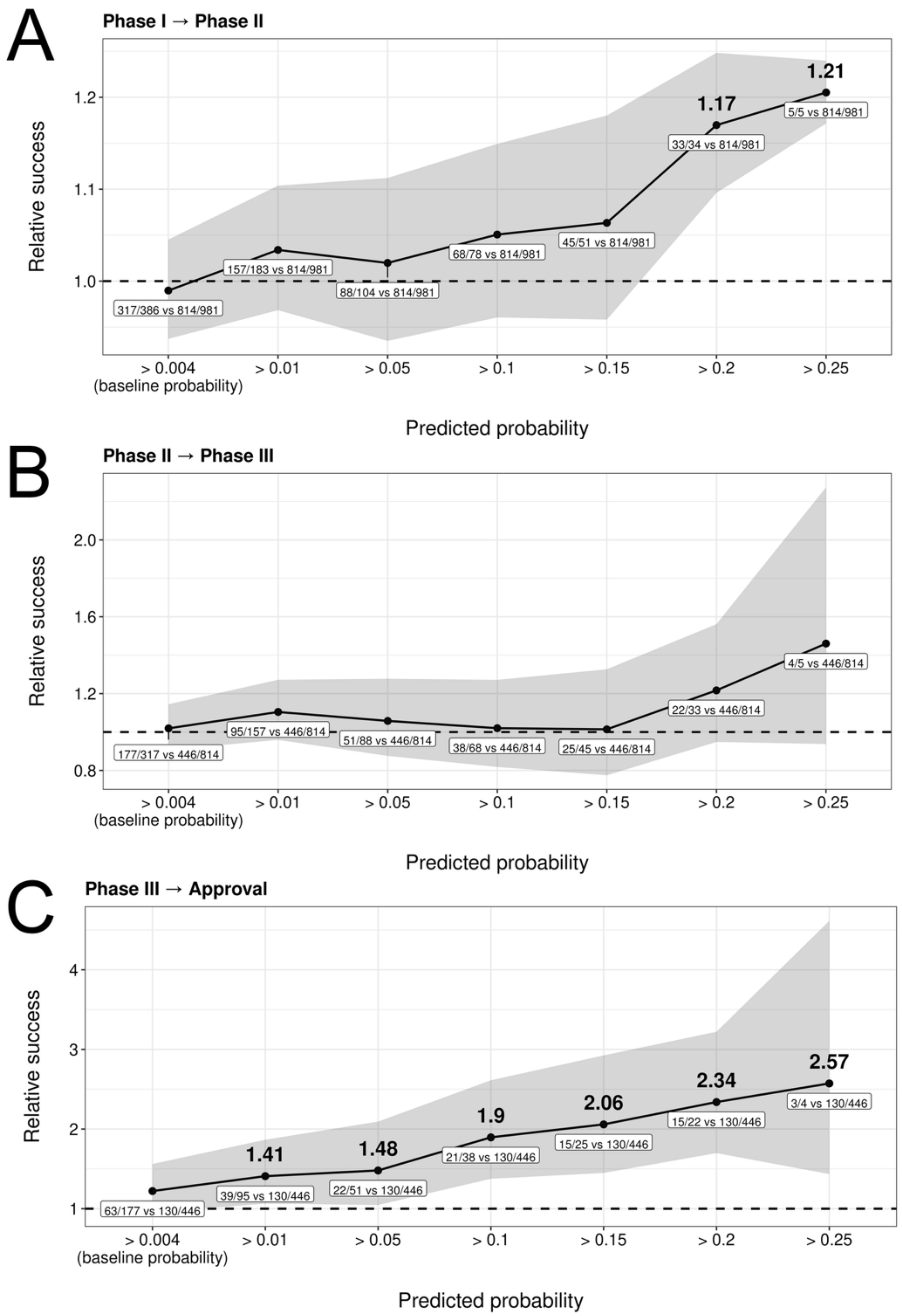


**Supplementary fig S8.** Relative likelihood (y-axis) of progressing from Phase X to Phase X + 1 for drug-disease pairs with predicted probability above a threshold (x-axis) compared to pairs below the baseline probability of being an indication in the test set (0.004). Values above each point indicate estimated relative success (fold-change; only when significantly different from 1). Values below points show the number of progressing drug-disease pairs relative to the total evaluated for pairs with predicted probability above the x-axis threshold versus those below the baseline probability of being indicated (0.004). Shaded regions indicate Katz 95% confidence intervals.

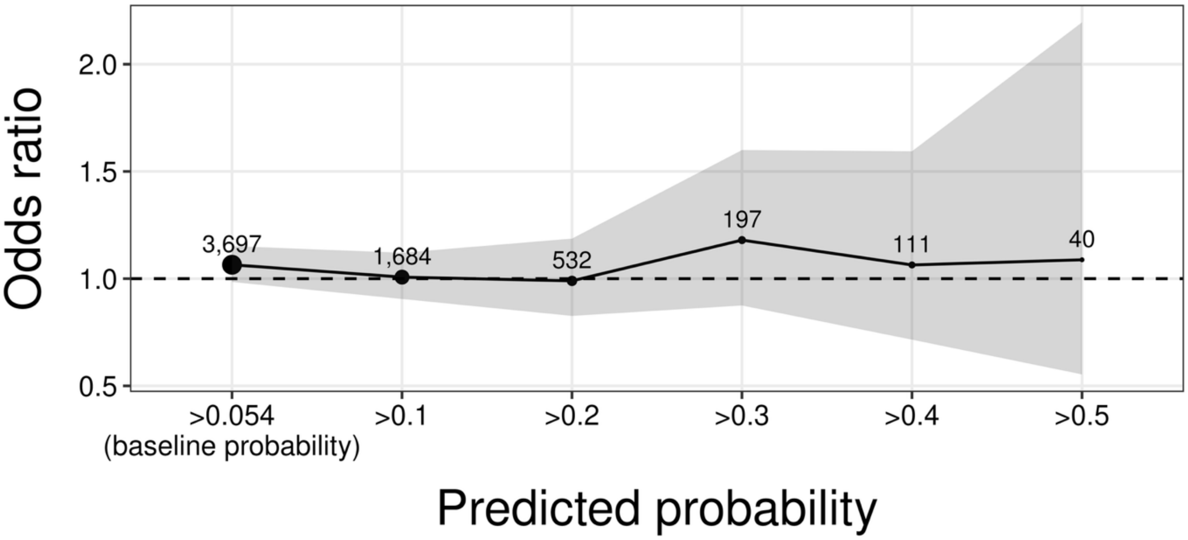


**Supplementary fig S9.** Odds ratio (y-axis) for drug-disease pairs with predicted probability greater than a given threshold (x-axis) being significantly reported in FAERS spontaneous side effect reports (offSIDES data) as side effect compared to those with predicted probability below the baseline probability of being a side effect in the test set (0.054). Shaded regions indicate 95% confidence intervals calculated by Fisher's exact test (two-sided). The total number of drug-disease pairs included in this analysis is 13,000 (drugs: 361; phenotypes: 159). The size of the points is proportional to the number of pairs with predicted probability greater than the x-axis threshold (actual number of pairs is printed above each point).

FAERS: FDA Adverse Event Reporting System

FDA: Food & Drug Administration

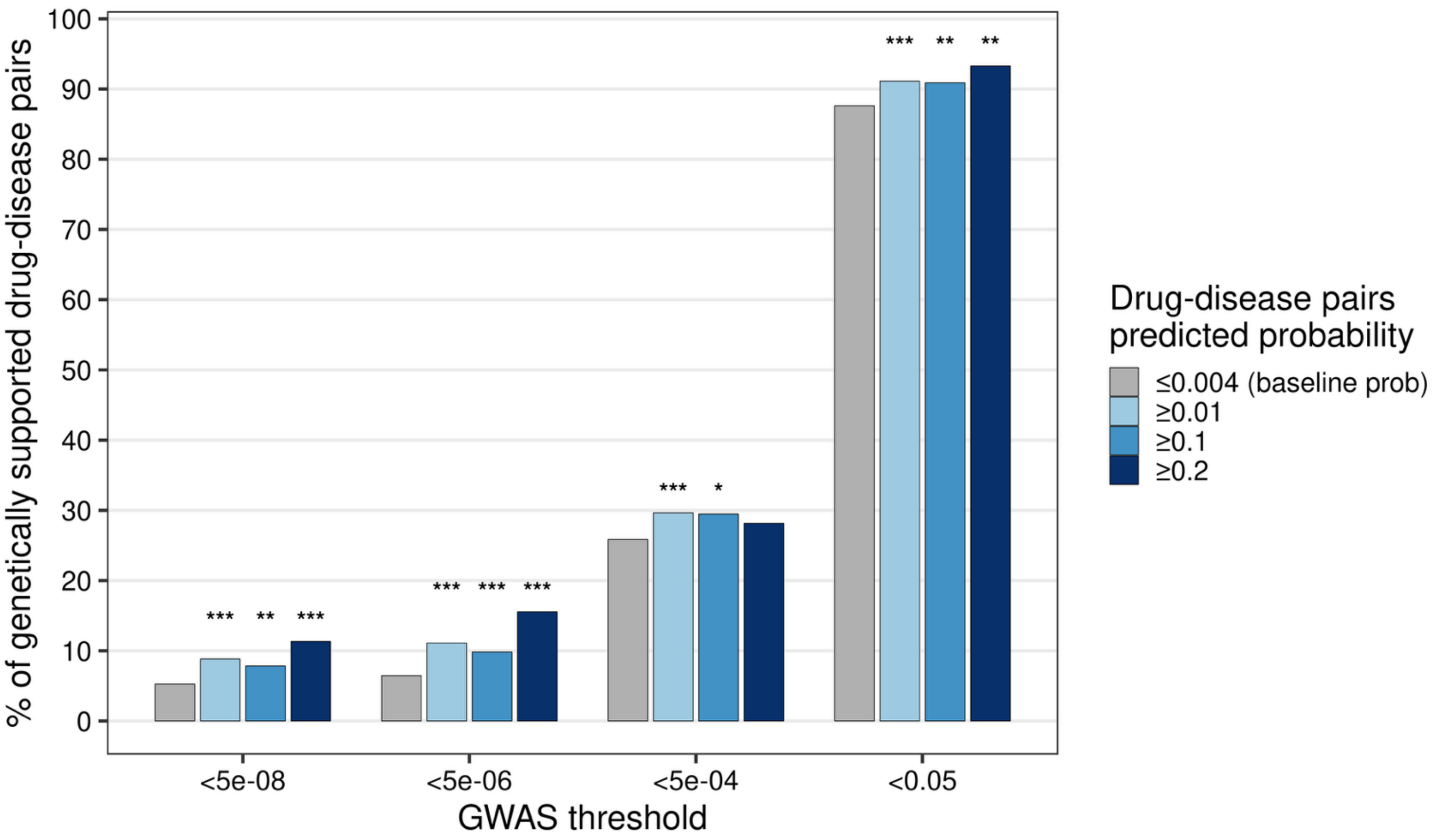


**Supplementary fig S10. Predicted drug-disease indication pairs involve drug gene-targets with disease-associated genetic variants, including subthreshold GWAS signals.** The y-axis shows the percentage of predicted drug-disease pairs in which the gene-targets of the drug contain at least one SNP with disease-specific GWAS p-value below a specified threshold (x-axis). Colors indicate groups of drug-disease pairs defined by predicted probability thresholds (legend). The gray group includes pairs with predicted probability below the baseline indication probability ($p=0.004$) and serves as the comparator group. Higher-probability groups are compared to this baseline group with respect to the proportion of pairs supported by genetic evidence (two proportions z-test, two-sided). Asterisks above the bars denote statistical significance (*: $p<0.05$; **: $p<0.01$; *** $p<0.001$). This analysis includes 99 diseases with available GWAS summary statistics on GWAS Catalog and their predicted drugs from our indications model trained on phenotypically dissimilar drug indication-disease pairs. We find SNPs located within the drug gene-targets genomic regions ±5kb using the R package BioMart. For each drug-disease pair, we keep the minimum p-value across SNPs within the drug's gene-targets.
